# Electrocorticography and stereo EEG provide distinct measures of brain connectivity: Implications for network models

**DOI:** 10.1101/2020.12.02.20242669

**Authors:** John M. Bernabei, T. Campbell Arnold, Preya Shah, Andrew Revell, Ian Z. Ong, Lohith G. Kini, Joel M. Stein, Russell T. Shinohara, Timothy H. Lucas, Kathryn A. Davis, Danielle S. Bassett, Brian Litt

## Abstract

Brain network models derived from graph theory have the potential to guide functional neurosurgery, and to improve rates of post-operative seizure freedom for patients with epilepsy. A barrier to applying these models clinically is that intracranial EEG electrode implantation strategies vary by center, region and country, from cortical grid & strip electrodes (ECoG), to purely stereotactic depth electrodes (SEEG), to a mixture of both. To determine whether models derived from one type of study are broadly applicable to others, we investigate the differences in brain networks mapped by ECoG and SEEG in a cohort of patients who underwent surgery for temporal lobe epilepsy and achieved a favorable outcome. We show that networks derived from ECoG and SEEG define distinct relationships between resected and spared tissue, which may be driven by sampling bias of temporal depth electrodes in patients with predominantly cortical grids. We propose a method of correcting for the effect of internodal distance that is specific to electrode type and explore how additional methods for spatially correcting for sampling bias affect network models. Ultimately, we find that smaller surgical targets tend to have lower connectivity with respect to the surrounding network, challenging notions that abnormal connectivity in the epileptogenic zone is typically high. Our findings suggest that effectively applying computational models to localize epileptic networks requires accounting for the effects of spatial sampling, particularly when analyzing both ECoG and SEEG recordings in the same cohort, and that future network studies of epilepsy surgery should also account for differences in focality between resection and ablation. We propose that these findings are broadly relevant to intracranial EEG network modeling in epilepsy and an important step in translating them clinically into patient care.

**Author summary:** Bernabei et al. report that electrocorticography and stereo EEG provide different quantifications of epileptogenic zone connectivity due to differences in electrode type and implant patterns. After correcting for sampling differences between modalities, they find that more focal forms of epilepsy surgery target regions of weaker connectivity compared to the remaining epileptic network.

## 1. Introduction

Intracranial electrode recordings from patients with medically-refractory epilepsy characterize the brain’s local function, widespread network organization, and guide surgical therapy. From the earliest days of intracranial EEG (iEEG), two major approaches have been used by clinicians for these purposes. In North America, Penfield’s use of subdural grid and strip electrodes for electrocorticography (ECoG) persists at many major centers now decades after its initial use^1,2^. Meanwhile, centers in France and Italy still favor Talaraich and Bancaud’s approach of using purely depth electrodes in stereoelectroencephalography (SEEG) pioneered at St. Anne’s Hospital in Paris^2,3^. Recently, many centers in the United States have begun to favor SEEG due to its superior risk profile and tolerability, though some centers continue to use ECoG for its superior cortical spatial coverage^4^. Unfortunately, many patients do not become seizure free after epilepsy surgery, regardless of implant technique. The reasons for poor outcomes are unclear, but it is likely in part because the interpretation of intracranial recordings is complex, subjective, and plagued by sampling uncertainty^5^. It is also difficult to determine where and how much of the epileptic network must be resected or ablated to fully prevent seizures, particularly in cases where there are no obvious lesions on MRI^6^. Validated, quantitative methods to guide epilepsy surgery could lead to a greater rate of seizure freedom and greater clinical benefit to patients.

Recent evidence supports the hypothesis that epilepsy arises from disordered connectivity^7,8^, and that mapping brain networks may aid in both selecting candidates for invasive treatment and identifying therapeutic targets for surgical resection, ablation or device implants^9^. In a brain network model, discrete ‘nodes’ exist either at the sensor-level for functional connectivity derived from iEEG signals, or at the atlas region-of-interest (ROI) level for structural connectivity derived from imaging^10^. Edges quantify the statistical relationships between nodes in functional connectivity approaches, or streamline count derived from DTI in structural connectivity. A variety of network methods are being explored to localize the epileptogenic zone from intracranial EEG data. Such approaches use interictal^11–13^ or ictal recordings^14–17^ and are derived from ECoG^11,12,14^ or SEEG^18,19^ to generate networks. These networks are analyzed using graph theory^11,14,20^ or neural mass models, which simulate seizure-like activity and probe the effects of different surgical interventions^12,17^. Intracranial EEG network models are also used to study networks activated during normal brain function, for example in recent studies probing cognition^21^ and attention^22^. The majority of these studies use patients implanted with ECoG, often supplemented with depth electrodes placed in the hippocampus. Unfortunately, because of the lack of standardization and difficulties in sharing iEEG data across centers, few studies test their methods in both ECoG and SEEG. In order to validate and translate network methods into clinical practice across centers, it is important to understand how these variations in electrode implantation impact estimate connectivity, subsequent network models and their clinical utility.

While networks derived from functional MRI and diffusion imaging easily generalize across patients and centers due to congruence in their full-brain spatial sampling, iEEG functional networks suffer from sparse sampling and implant heterogeneity. Still, the problem of spatial sampling bias which affects iEEG networks^23^ may be offset by: (1) the superior spatiotemporal resolution of iEEG in implanted regions, compared to functional neuroimaging and (2) clinical experience that associates particular patterns in the EEG with typical onset regions, though it can sometimes be difficult to tell if these patterns are the result of seizure generation or spread. To better translate network models into patient care, we must better understand the extent of bias or sensitivity introduced by electrode implantation strategy, in this case ECoG versus SEEG, and its effect on network models. We must then either change implant strategy or develop computational methods to correct for this effect. It is important to note that tradition in specific centers is not the only thing that guides choice of electrode implantation strategy. Other issues, such as the need for stimulation mapping, characteristics of a lesion such as its type and location, ictal semiology and suspected clinical syndrome, as well surgeon and epileptologist experience and training may also factor into approach and electrode choice^24^.

There are many differences in implantation strategy between ECoG and SEEG that arise from the electrode hardware itself. Patients implanted with a large ECoG grid will have regular spacing between contacts in the same electrode (e.g. an 8×8 contact grid), supplemented with a few additional strip and depth electrodes in other regions, as needed. The implantation strategy is much more heterogeneous in SEEG. This heterogeneity can manifest itself in the following ways: (i) a wide range in the number of depth electrodes & electrode contacts from center to center, (ii) different spatial orientations and density of depth electrode implantation, (iii) different assortments of anatomical targets, electrode spacing and (iv) different levels of implant bilaterality^4^. Thus, translating network models into clinical care faces the challenge not only of resolving differences between ECoG and SEEG approaches, but also a high variability in SEEG approaches from center to center. It is unclear whether these distinct properties of electrodes and implant strategy preferentially degrade network representations derived from one approach vs the other, and whether they preclude using the same analysis for networks derived from SEEG and ECoG.

In this study, we explore the effect of implant strategy in a retrospective cohort of patients with drug-resistant temporal lobe epilepsy who were evaluated with either ECoG or SEEG for invasive treatment. We hypothesize the following: (i) ECoG and SEEG networks have distinct properties due to different patterns of spatial sampling, and (ii) differences in network properties between ECoG and SEEG will impact the observed relationship between resected and spared tissue. We aim for our findings to help translate personalized network models of epilepsy into clinical practice, and to inform other applications of intracranial EEG connectivity analysis.

## 2. Methods

### 2.1 Patient data acquisition

We retrospectively analyzed a data set consisting of 33 patients who underwent intracranial recording during evaluation for epilepsy surgery at the Hospital of the University of Pennsylvania (HUP). Sixteen of these patients had implants with grid, strip, and a small number of depth electrodes, while the remaining patients had only stereotactically-placed depth electrodes. In this study we refer to cortical-predominant patients as the ‘ECoG’ group while patients with only depth electrodes constitute the ‘SEEG’ group. All patients underwent either resection or laser ablation after electrode explant, however in subsequent sections we use the term ‘resected’ tissue to include ablation patients as well. We chose only patients that achieved good outcome, assessed at 6 months post-operatively, to maximize the likelihood that tissue removed in surgery contained the epileptogenic zone. Table 1 lists subject demographics and characteristics of therapy and electrode implants, while Supplementary Table 1 contains the same information on a per-patient basis. All subjects provided consent to have their full-length intracranial EEG recordings and anonymized imaging and metadata publicly released on the ieeg.org portal, an open-source online repository for electrophysiologic studies^25,26^.

**Table 1:** Clinical and demographic information. We analyzed a retrospective cohort of 33 patients with drug-resistant epilepsy who underwent surgery of the temporal lobe and achieved seizure freedom at 6 months post-operatively. Abbreviations: GM: grey matter. ^a^ = Fisher’s exact test; ^b^ = Wilcoxon rank-sum test.

|  | ECoG | SEEG | p-value |
| --- | --- | --- | --- |
| Total number of subjects | 16 | 17 |  |
| Number of female subjects | 10 | 8 | 0.49<sup>a</sup> |
| MRI* |  |  | 0.75<sup>a</sup> |
| Lesional | 8 | 10 |  |
| Non-Lesional | 8 | 8 |  |
| Type of surgery |  |  | **0.0033<sup>a</sup>** |
| Resection | 14 | 7 |  |
| Laser ablation | 2 | 11 |  |
| Node counts |  |  |  |
| Total GM contacts |  |  |  |
| Mean ± std. dev. | 92.1 ± 21.2 | 88.6 ± 34.7 | 0.72<sup>b</sup> |
| Depth GM contacts |  |  |  |
| Mean ± std. dev. | 10.9 ± 9.3 | 88.6 ± 35.4 | **1.5e-6<sup>b</sup>** |
| Total GM resected / ablated |  |  |  |
| Mean ± std. dev. | 16.9 ± 14.0 | 9.1 ± 6.0 | 0.08<sup>b</sup> |
| Depth GM resected / ablated |  |  |  |
| Mean ± std. dev. | 4.1 ± 4.8 | 9.1 ± 6.0 | **0.01<sup>b</sup>** |

Each patient underwent a standard epilepsy imaging protocol including pre-implant MRI, post implant CT & MRI, and post-resection MRI. We have previously described our method for localizing electrode locations in detail^14,27^, and briefly summarize them in Figure 1. Post-iEEG-implant MRI (Figure 1D) was registered to pre-implant MRI (Figure 1C) using ANTs^28^ and electrodes are segmented to derive their coordinates using ITK-SNAP^29^. Any electrode contacts with centroids outside of the brain in the native MRI space were eliminated. We then non-linearly registered the pre-implant MRI into Montreal Neurological Institute (MNI) space for use with neuroimaging atlases, and visually inspected results for accuracy in each subject. We chose a 90 region AAL atlas^30^ to assign each electrode contact location a brain region of interest (ROI). Any electrodes in white matter with centroids not overlapping with any atlas region were eliminated. Finally, we used a semi-automated algorithm previously described and validated^14^ to perform resection and ablation zone segmentations, which allow the electrode contacts targeted by surgery to be determined.

**Figure 1.**
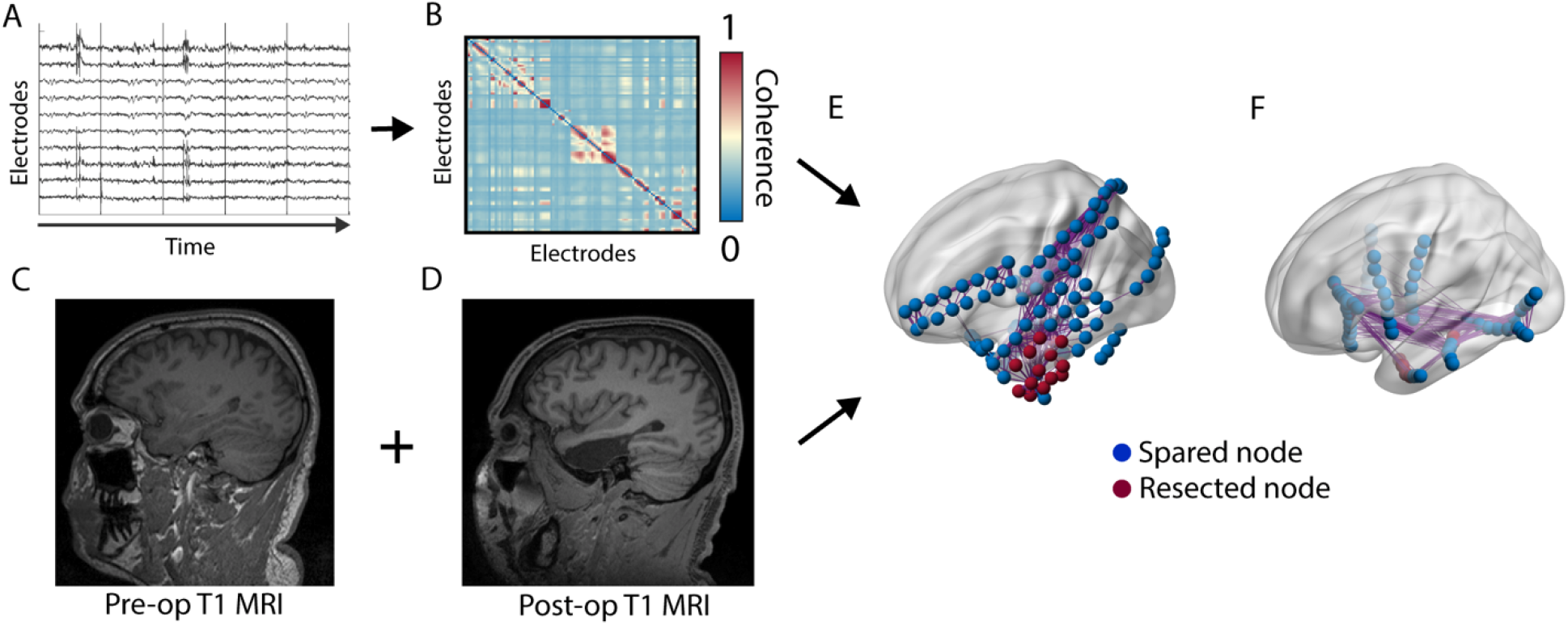
Imaging and network methods: (A) We use artifact-free clips of interictal iEEG to calculate (B) mean adjacency matrices using multitaper coherence. (C) Pre-operative and (D) post-operative T1 weighted MRI are used to segment the resection cavity which is used to determine resected nodes. (E) Together, we construct networks with the resected nodes determined in ECoG. (F) SEEG implantations using only depth electrodes appear distinct even for similar anatomic targets.

### 2.2 Connectivity calculation

We calculated functional connectivity using a pipeline that we have previously described and validated^11,14,16^. We randomly selected an interictal segment 1 hour in length for each patient, occurring at least 1 hour away from clinically-annotated seizures. We divided the interictal epoch into one-second intervals (Figure 1A) and for each window calculated connectivity using coherence in the beta (15-25 Hz) and low-gamma (30-40 Hz) bands as well as using broadband cross correlation (after applying 5-115 Hz bandpass and 60 Hz notch filters) as they previously have yielded significant results in interictal network studies in ECoG subjects^11^. We computed the median of each edge over time to obtain a single adjacency matrix for each patient (Figure 1B). Together with the results of our imaging pipeline, this process yielded networks in which each node is either resected/ablated or spared in both patients with ECoG (Figure 1E) and SEEG (Figure 1F).

### 2.3 Network methods

To probe how network structure differs between ECoG and SEEG we detected communities using modularity maximization, which labels nodes so that each community consists of nodes that are more connected to each other and relatively less connected to all other nodes outside of their community^31^. We used a Louvain-like method^32^ to maximize modularity, which is represented by 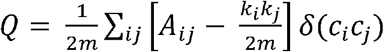, where *A*_*ij*_ is link between nodes *i* and *j, k* are edge weights, *m* is the sum of all edge weights in the graph, and δ is the Kronecker delta function. To compare community structure across patients, we computed the participation coefficient^33^ which measures the ratio of a node’s connectivity strength external versus internal to its module. Averaging participation coefficient across nodes within each patient yielded an estimate of whether networks are (i) integrated, with high connectivity between modules, or (ii) segregated, with lower connectivity between modules.

To illustrate the importance of differences in the way ECoG and SEEG represent epileptic networks in-vivo, we compared the ability of connectivity derived from these modalities to distinguish resected and spared tissue. We chose the simple network metric of node strength, computed as the sum of all edge weights connecting it to all other nodes and is computed as 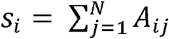 in which *s*_*i*_ is the strength of node *i, A*_*ij*_ is the adjacency matrix element containing the edge weight between node *j* and node *i*, and *N* is the number of nodes. Our group and others have previously demonstrated that high node strength localizes the epileptogenic zone and predicts surgical outcome in patients implanted with ECoG^11,20^, however its translatability to SEEG is not well-established.

### 2.4 Statistical Approach

In our comparisons of network properties between ECoG and SEEG groups we primarily used nonparametric statistical tests such as the Wilxocon rank-sum test, as they do not assume that data are normally distributed. To assess the ability for node strength to detect epileptogenic regions, we used a metric known as the distinguishability statistic (D_rs_) which quantifies the area under the receiver operating characteristic curve for classifying nodes as either resected or non-resected^34^. The quantity D_rs_ has been previously studied and validated for its ability to quantify whether networks have sufficient information to determine resected or non-resected regions^34,35^. This value is calculated as the normalized U-statistic, and ranges from 0-1. In our study, a value of 1 implies that all EZ nodes are lower in strength than all non-resected nodes; a value of 0 means that all EZ nodes are stronger than all non-resected nodes; and a value of 0.5 implies that node strength is unable to distinguish between resected and spared nodes.

### 2.5 Data availability

One of our primary goals is to aid in the translation of epilepsy network models into clinical practice. To this end, we shared all raw intracranial EEG and imaging data for HUP patients at iEEG.org, a free cloud sharing platform for electrophysiological data. Each subject’s recordings are associated with the ID listed in Supplementary Table 1 and can be accessed through the web interface or the open-source iEEG.org MATLAB & python toolboxes. The code for calculating adjacency matrices is available at GitHub.com/Akhambhati/echobase, processed adjacency matrices, and code for comparing networks between ECoG and SEEG is hosted at GitHub.com/jbernabei/ecog_vs_seeg.

## 3. Results

Here we compared networks mapped by ECoG and SEEG in a cohort of temporal lobe epilepsy patients. We aimed to describe how each implant approach is biased towards distinct network properties. We then compared how these distinct network properties affected how well connectivity could distinguish epileptogenic and non-epileptogenic regions with the ultimate goal of improved surgical planning.

### 3.1 Anatomical sampling is similar between modalities

We first asked what differences in the location and extent of anatomic sampling exist between ECoG and SEEG. After quantifying the top 15 anatomical targets implanted by each approach (Supplemental Figure 1A), we found a similar distribution of electrode contacts with temporal gyri, hippocampus, and inferior frontal gyri highly sampled. While the total number of implanted nodes was higher in SEEG compared to ECoG (120.6 ± 41.5 vs. 94.5 ± 22.3, rank-sum p < 0.05), many depth electrodes localized to white matter and ultimately the number of nodes in grey matter (GM) across ECoG and SEEG was similar (92.1 ± 21.2 vs. 88.6 ± 34.7, rank-sum p = 0.7, Table 1). Ensuring similar node count was critical for comparing networks and thus for all subsequent analyses we considered only GM nodes. We observed a slight bias of ECoG to favor ipsilateral sampling with more nodes than SEEG implanted in the same hemisphere as the resection zone (Supplemental Figure 1B, rank-sum test p = 0.02). The median number of contralateral nodes was higher in SEEG than ECoG (25 vs. 4), however this did not reach statistical significance (Supplemental Figure 1C, rank-sum p = 0.06). Despite the modest differences in hemispheric differences in GM nodes, we observed similar median internodal distances between ECoG and SEEG (Figure S1D, S1E, rank-sum p = 0.2). Overall, the targets sampled by ECoG and SEEG for patients with temporal lobe epilepsy were similar, implying that differences in anatomy and internodal distance alone would not primarily drive any subsequent differences in network models.

### 3.2 Differences in mapping resected versus spared tissue

Although ECoG and SEEG sample from similar brain regions, they may not represent the epileptogenic regions similarly from a network perspective. We aimed to gauge the ability of each implant strategy to distinguish resected and spared tissue using the distinguishability statistic (D_rs_). This value is high when resected nodes are weaker than the spared network and low when they are stronger (Figure 2A). Across our cohort, ECoG patients tended to have a low D_rs_ value while SEEG had higher and more variable values (rank-sum test, p < 0.01), which was unexpected given that all patients had temporal lobe epilepsy and achieved good surgical outcome (Figure 2B). We then sought to determine whether the difference network relationship between resected and spared tissue could result from the frequent placement of depth temporal depth electrodes in ECoG subjects (Figure 2C). In these patients we found that resected nodes from surface electrodes had higher normalized strength than non-resected surface electrodes (rank-sum test, p < 0.01), however resected depth electrodes were not higher in strength than non-resected depth electrodes (rank-sum test, p = 0.1). Furthermore, non-resected depth electrodes were higher in strength than non-resected surface electrodes (rank-sum test, p < 0.01), and resected depth electrodes were higher in strength than resected surface electrodes (rank-sum test, p < 0.01). These findings, and the sizable proportion of resected depth electrode contacts in ECoG could account for the observed difference in D_rs_ values between the two implantation strategies.

**Figure 2:**
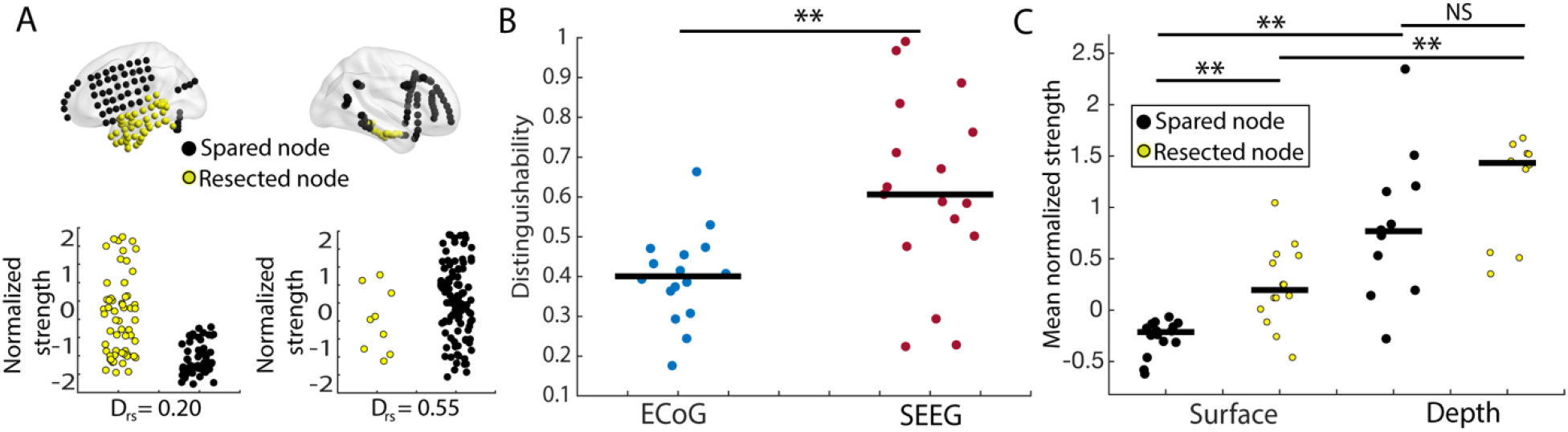
Network localization. (A) Distinguishability statistic calculated for an ECoG patient (left) and a SEEG patient (right). In cases where resected node strength is higher than the remaining network on average, D_rs_ will have a low value, in cases where resected node strength is lower than the remaining network D_rs_ will be high. A D_rs_ value of 0.5 means that node strength cannot distinguish resected and spared tissue (B) In networks of grey matter nodes, D_rs_ of resected and spared tissue is higher in SEEG compared to ECoG (rank-sum test, p = 0.0026). (C) In patients with ECoG we found resected nodes from surface electrodes to be higher in strength than non-resected surface electrodes (rank-sum test, p = 0.0065). Non-resected depth electrodes were higher in strength than non-resected surface electrodes (rank-sum test, p = 0.0013). Resected depth electrodes were higher in strength than resected surface electrodes (rank-sum test, p = 0.0031). Resected depth electrodes were not higher in strength than non-resected depth electrodes (rank-sum test, p = 0.14). ** = p < 0.01.

### 3.3 Distinct network properties between ECoG and SEEG

We sought next to determine whether we could adequately correct for our findings of D_rs_ differences between SEEG and ECoG by regressing for internodal distance in an electrode-specific manner. We fit a nonlinear regression model to ECoG and SEEG separately (Figure 3A) using a rational polynomial (rat11 in MATLAB) which has been previously validated in interictal network analysis for epilepsy^35^. Within ECoG we also used different models for depth-depth connections, surface-depth connections, and surface-surface connections. Even after correcting for internodal distance, network heterogeneity was higher in SEEG (Figure 3B), represented by a higher standard deviation of edge weight residuals across the network (rank-sum test, p < 0.01). Furthermore, we calculated modularity in distance-corrected networks (Figure 3C) and found a higher median participation coefficient in SEEG representing higher network integration (rank-sum test, p < 0.01). Analogous results for broadband cross – correlation and low – gamma coherence are found in Supplementary Figure 3. These results indicate that despite accounting for internodal distance and the effects of distinct electrode types on connectivity, differences in global network properties remain.

**Figure 3:**
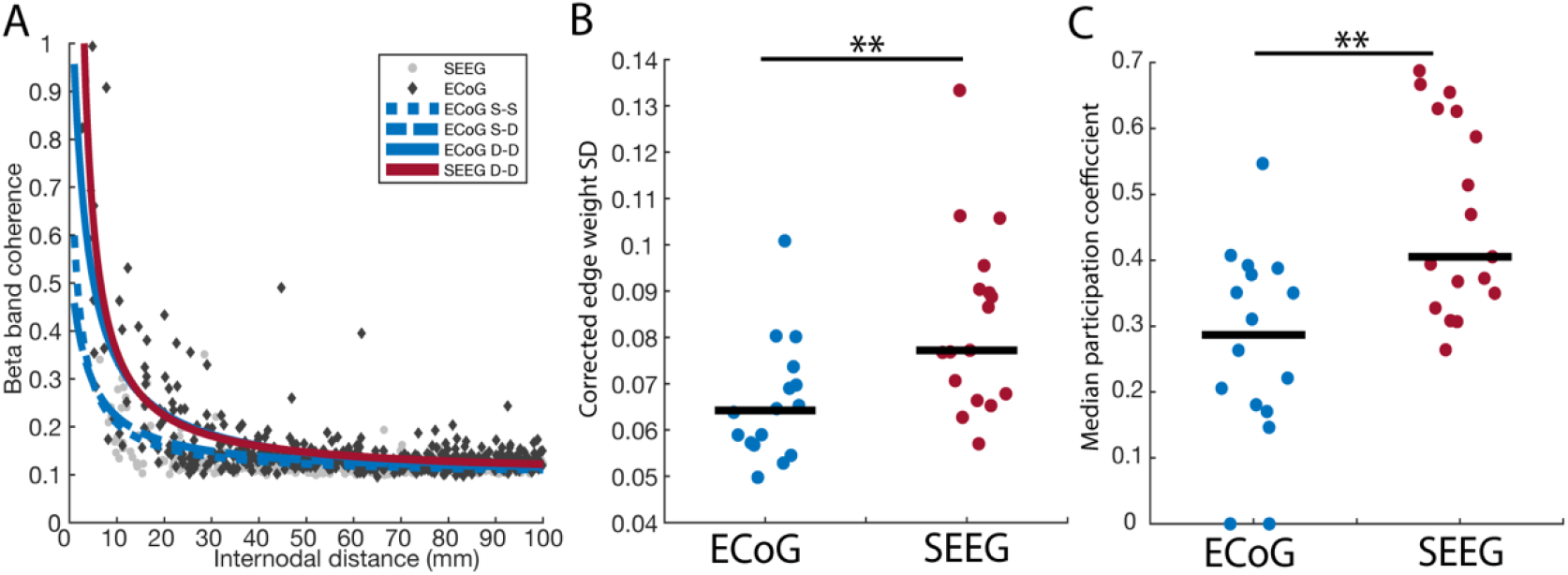
Global network structure is impacted by sampling differences between ECoG and SEEG. (A) We fit a nonlinear regression model to ECoG surface – surface (dotted blue line), surface – depth (dashed blue line), and depth – depth connections (solid blue line), as well as SEEG depth – depth connections (solid red line). (B) After correcting for internodal distance, the standard deviation of edge weights remained higher in SEEG versus ECoG (rank-sum test p = 0.0052). (C) After correcting for internodal distance, the median participation coefficient remained higher in SEEG versus ECoG (rank-sum test p = 0.0018). Abbreviations: S-S: surface – surface, S-D: surface – depth, D-D: depth – depth, SD: standard deviation, ** = p < 0.01.

### 3.4 Modifying ECoG and SEEG networks to correct for sampling bias

We finally asked how our internodal distance correction would affect resection zone distinguishability in ECoG and SEEG, and whether we could correct for any remaining differences in network localization by simplifying initial networks to reduce sampling differences. Due to the different balance of ipsilateral and contralateral nodes in ECoG and SEEG (Supplemental Figure 1B & C) and the distinct connectivity of intra-vs inter-hemispheric edges, we hypothesized that eliminating nodes contralateral to the resection zone of distance-corrected networks could improve localization. Based on the distinct modular structure of ECoG and SEEG, we additionally hypothesized that averaging all edges between pairs of brain regions and thus reducing nodes from representing electrode contacts to representing atlas-level regions of interest (ROI) could correct for different balances of connectivity within and between modules. We performed each of these steps for all ECoG and SEEG patients (Figure 4A).

**Figure 4:**
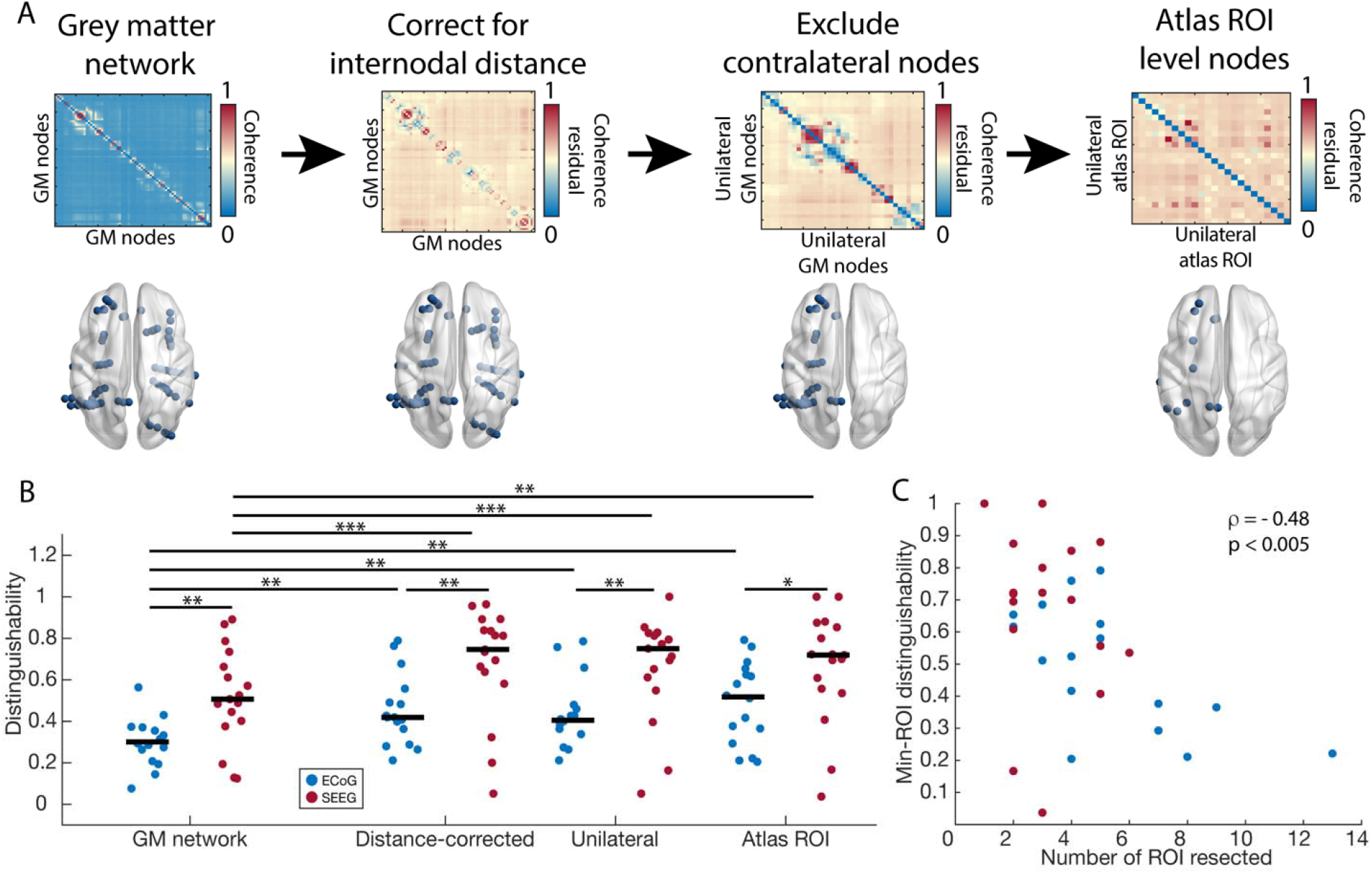
ECoG and SEEG have distinct representations of the epileptogenic zone. (A) We used three approaches to modifying networks to probe sampling differences between ECoG and SEEG and their effect on distinguishing resected and spared tissue. We corrected for the effects of internodal distance (DC). Then, we eliminated nodes contralateral to the resection zone (UL). Finally, we averaged edges between pairs of brain regions to have a single node per atlas-level region of interest (AR). (B) We compared the effect of correcting for internodal distance to unilateral to atlas ROI representations. For each condition, SEEG networks had a higher D_rs_ value than ECoG. Each condition in ECoG and SEEG also had a higher D_rs_ value than not accounting for internodal distance. GM ECoG vs GM SEEG, as in Fig. 2B: (rank-sum test p = 0.0065), DC ECoG vs DC SEEG: (rank-sum test p = 0.0059), UL ECoG vs UL SEEG: (rank-sum test p = 0.0047), MR ECoG vs MR SEEG: (rank-sum test p = 0.022). DC/UL/AR ECoG vs GM ECoG: (sign-rank test p = 0.0027/0.0061/0.0011), DC/UL/AR SEEG vs GM SEEG: (sign-rank test p = 0.0006/0.0008/0.0031). (C) D_rs_ values of min-ROI networks were negatively correlated with the number of ROI that contained electrode contacts in the resection zone (Pearson correlation ρ = −0.48, p < 0.005).

We then calculated D_rs_ for each of the three modifications (i) correction for internodal distance (DC), (ii) using only unilateral nodes ipsilateral to the resection zone (UL), and (iii) atlas-level ROI (AR) (Figure 4B). For each condition, SEEG networks had a higher D_rs_ value than ECoG: GM ECoG vs. GM SEEG, as in Fig. 2B: rank-sum test p < 0.01, DC ECoG vs. DC SEEG: rank-sum test p < 0.01, UL ECoG vs. UL SEEG: rank-sum test p < 0.01, AR ECoG vs. AR SEEG: rank-sum test p = 0.02. Each condition in ECoG and SEEG also had a higher D_rs_ value than in uncorrected networks (DC / UL / AR ECoG vs GM ECoG: sign-rank test p < 0.01 for ech, DC / UL / AR SEEG vs GM SEEG: sign-rank test p < 0.001 / 0.001 / 0.01). Analogous results for broadband cross – correlation and low – gamma coherence are found in Supplementary Figure 2. The atlas-ROI representation had the highest median distinguishability in ECoG compared to the base network, however SEEG patients had higher D_rs_ values compared to ECoG even for this condition. As ECoG and SEEG networks with distance-corrected, unilateral, ROI-level nodes should be as similar as possible, we hypothesized that remaining differences were a result of differences in the extent of surgical intervention between groups (Table 1). Indeed, across ECoG and SEEG, we found a strong, negative correlation between D_rs_ values and the number of atlas ROI nodes resected (Pearson correlation rho = −0.48, p < 0.005, Figure 4C). This finding suggests that the network relationship between resected and spared tissue depends on the focality of the surgical approach.

## 4. Discussion

Understanding the sampling differences of different implant approaches is critically important when applying network models to interpret iEEG data. Here, we showed how unique characteristics of ECoG and SEEG sampling result in distinct properties of derived networks despite similar clinical targets. Node strength, a frequently studied network metric in epilepsy, had an unpredictable relationship between resected and spared tissue, and accounting for internodal distance & electrode type still resulted in distinct network properties. We also establish that these general patterns are present in different frequency bands and in both coherence and correlation measures of functional connectivity. Finally, we provided several methods to partially mitigate the effects of sampling bias introduced by implantation strategy on network models, and showed that remaining differences are associated with the focality of the subsequent resection or ablation.

Our study adds to the growing body of literature on methodological considerations for applying network models clinically^23,35^. From these studies, we recognize that a major challenge in applying network models to the epileptic brain is determining whether observed patterns in brain activity and therefore network structure truly capture the phenomena of interest, and to what extent they arise from sampling artifact. To this end, we must acknowledge that the sampling bias in both ECoG and SEEG is distinct, and that subtle differences in the arrangement of electrodes can determine whether connectivity can accurately uncover true pathology. Thus, sampling bias is more complex than just whether or not a particular target was sampled, and we must take special care to ensure that models are not biased by the locations of electrodes selected by physicians to simply confirm *a priori* suspicions present before implant.

While the finding that ECoG and SEEG have distinct network connectivity patterns in resected versus spared tissue is significant, it may in part reflect conceptual differences underlying these implant strategies. ECoG attempts to map the boundaries of epileptogenic cortical regions by assessing seizures and interictal activity^3^, while SEEG focuses on ‘electro-anatomo-clinical’ correlations^3^, in which broader network mapping and the relationship of anatomical spread to seizure semiology is important. For these reasons, as well as the typical use of a single electrode type & geometry, that SEEG may be superior from a network perspective as the inherent conceptualization of the modality takes the network approach in mind^36^. In particular, reducing networks to atlas ROI nodes as we present here may be an appealing approach for this type of mapping in the future, since these regions and their connections correspond to anatomically relevant and interpretable structures. Such an approach could also facilitate the integration of findings from intracranial EEG networks with studies of quantitative imaging such as fMRI and DTI which typically use atlas ROI nodes, or through the use of intracranial EEG atlases for the prediction of missing information^37,38^.

While others have reported differences in connectivity values between depth electrodes and surface electrodes^39^, the potential scientific and clinical relevance of network differences that arise as a result of these are significant. It is likely that this finding underlies the results of Figure 2B, that in uncorrected networks of ECoG which often include temporal depths, connectivity of resected tissue is relatively strong whereas in SEEG it is variable. Indeed, in cases of suspected temporal lobe epilepsy mapped by ECoG, the chance that depth electrodes will capture the seizure onset zone is high. Furthermore, the different physical size and cylindrical shape of each electrode contact in SEEG compared to ECoG records local fields from different types of neural populations which could have distinct coherence values. Overall, this observation is likely fixed by performing separate internodal distance corrections for depth and cortical electrodes, which adds to the literature that regressing for internodal distance improves outcome prediction^35^.

The results of our sampling correction process (Figure 4) reveal interesting aspects of sampling differences between ECoG and SEEG. The finding that eliminating nodes contralateral to the resection zone doesn’t significantly change localization from bilateral distance-corrected networks, implies that this issue is not a major factor driving why certain models may succeed or fail in some patients. Indeed, most subjects do not have symmetric implants but rather have a bias towards the hemisphere with the most clinical correlates. Contralateral electrodes are often placed to address clinical hypotheses of lateralization, and due to their relative isolation from the bulk of electrodes, it is possible that they are already outliers in the network and do not contribute highly to the outcome of the distinguishability statistic. On the other hand, if the true epileptogenic zone is in the hemisphere with fewer electrodes, network models may struggle with localizing it. Furthermore, our finding that averaging edges between pairs of brain regions to convert nodes to atlas ROIs maintains similar performance implies that network models may not need dense sampling within regions, but instead may benefit from sampling inter-regional connections.

Another important consideration to the application of network models is the issue of surgical focality which may differ significantly between resection and ablation patients. While many previous studies focus on resection patients, which may extend to natural anatomic boundaries therefore removing additional, non-epileptogenic tissue, a large part of our cohort underwent ablation in which lesions are relatively small and more specific to the epileptogenic zone. Indeed, others have found that a large number of nodes within and outside of the resection zone is important for accurate outcome prediction from interictal connectivity, which may be impossible for ablation patients. In this context, our finding of a negative correlation between D_rs_ and number of regions targeted may imply that the truest representation of the epileptogenic zone is of low node strength relative to the rest of the brain. This notion is supported by studies demonstrating cellular loss in these regions, particularly the hippocampus, in temporal lobe epilepsy^40,41^. As minimally-invasive approaches such as laser ablation become more common, it is important that our notions of network abnormalities and methods of localizing the epileptogenic zone and outcome prediction do not rely too heavily on findings from resection patients alone.

Despite its encouraging results in illuminating the differences between ECoG and SEEG networks, our study has several key limitations. We focused our analysis on node strength, one of the simplest graph theory metrics that has been studied frequently in epilepsy, so this measure may not fully represent the complexity of abnormal networks in this disorder. However, given that node strength is among the least sensitive network metrics to sampling bias^23^, we felt that it was a reasonable choice to compare these approaches. Furthermore, It is well known that node strength is correlated with other network centrality metrics^42^ and even certain phenomenological network models which employ dynamical systems linked by functional connectivity^12^. Thus, many of the principles we highlight here may be broadly generalizable to other network studies in epilepsy, and future work should account for sampling bias where possible. Another limitation is our consideration of a single 1-hour iEEG segment, which does not capture the variability in interictal activity and thus connectivity which is known to follow circadian^39^ and slower timescales^43^. However, sleep-wake cycles may be interrupted and difficult to estimate in the epilepsy monitoring unit when sleep deprivation and medication withdrawal are common, and hospital admissions are too short to capture predominant multi-day cycles which are close to a month long in many patients. A final limitation is our analysis of only temporal lobe epilepsy patients. We chose this cohort to minimize variability within and across groups and because they represent the largest number of patients at our center. However, distinct patterns of sampling bias may exist in extratemporal epilepsies. Future work should address whether the relationship of connectivity in other anatomical locations of the epileptogenic has a similar pattern to temporal lobe epilepsy, and whether these patterns are also different in ECoG versus SEEG.

Ultimately, clinical judgement of risk and reward will drive the choice between ECoG and SEEG for individual patients. While the path of clinical translation for network models is complicated by inherent sampling biases across techniques and varying surgical practice among institutions, we believe it is vital to compare and contrast studies of ECoG and SEEG and recognizing that each provides a distinct view of the brain’s underlying connectivity but that neither are ‘correct’. Finally, we believe that carefully understanding the sampling properties of networks mapped by intracranial EEG can extend the use of graph theory to broader problems in translational human neuroscience.

## Data Availability

One of our primary goals is to aid in the translation of epilepsy network models into clinical practice. To this end, we share all raw intracranial EEG and imaging data for HUP patients at iEEG.org, a free cloud sharing platform for electrophysiological data in the folder HUP_Intracranial_Data. The code for calculating adjacency matrices is available at GitHub.com/Akhambhati/echobase, and the metadata, processed adjacency matrices, and code for comparing networks between ECoG and SEEG is hosted at GitHub.com/jbernabei/ecog_vs_seeg.

https://www.ieeg.org/

## 5. Acknowledgements

J.B. acknowledges funding from NIH 6T32NS091006. B.L. acknowledges funding from the Pennsylvania Tobacco Fund, NINDS R01: NS099348, NIH DP1: NS122038, the Mirowski Family Foundation, Jonathan Rothberg, and Neil and Barbara Smit. K.A.D acknowledges funding from NINDS K23: NS092973 and the Thornton Foundation.

## References

1. Lüders, H. O., Najm, I., Nair, D., Widdess-Walsh, P. & Bingman, W. Definition and localization of the epileptogenic zone The epileptogenic zone: general principles. Epileptic Disord. 8, S1–9 (2006).

2. Reif, P. S., Strzelczyk, A. & Rosenow, F. The history of invasive EEG evaluation in epilepsy patients. Seizure 41, 191–195 (2016).

3. Jehi, L. The Epileptogenic Zone: Concept and Definition. Epilepsy Curr. 18, 12–16 (2018).

4. Youngerman, B. E., Khan, F. A. & McKhann, G. M. Stereoelectroencephalography in epilepsy, cognitive neurophysiology, and psychiatric disease: Safety, efficacy, and place in therapy. Neuropsychiatric Disease and Treatment 15, 1701–1716 (2019).

5. Parvizi, J. & Kastner, S. Human Intracranial EEG: Promises and Limitations. Nat. Neurosci. 21, 474–483 (2018).

6. Téllez-Zenteno, J. F., Ronquillo, L. H., Moien-Afshari, F. & Wiebe, S. Surgical outcomes in lesional and non-lesional epilepsy: A systematic review and meta-analysis. Epilepsy Res. 89, 310–318 (2010).

7. Neuroscientist, C., Kramer, M. A. & Cash, S. S. Overview of Epilepsy and Seizures, Rhythms, and Networks Different Kinds of Seizures and Implications for Spatial Characterization of Epilepsy Epilepsy as a Disorder of Cortical Network Organization. Neurosci. 18, 360–372 (2012).

8. Bernhardt, B. C., Bonilha, L. & Gross, D. W. Network analysis for a network disorder: The emerging role of graph theory in the study of epilepsy. Epilepsy and Behavior 50, (2015).

9. Scheid, B. H. et al. Synchronizability predicts effective responsive neurostimulation for epilepsy prior to treatment. medRxiv 2021.02.05.21250075 (2021). doi:10.1101/2021.02.05.21250075

10. Bassett, D. S., Zurn, P. & Gold, J. I. On the nature and use of models in network neuroscience. Nat. Rev. Neurosci. 19, 566–578 (2018).

11. Shah, P. et al. High interictal connectivity within the resection zone is associated with favorable post-surgical outcomes in focal epilepsy patients. NeuroImage Clin. 23, 101908 (2019).

12. Sinha, N. et al. Predicting neurosurgical outcomes in focal epilepsy patients using computational modelling. Brain 140, 319–332 (2017).

13. Lagarde, S. et al. Interictal stereotactic-EEG functional connectivity in refractory focal epilepsies. Brain 141, 2966–2980 (2018).

14. Kini, L. G. et al. Virtual resection predicts surgical outcome for drug-resistant epilepsy. Brain 142, 3892–3905 (2019).

15. Khambhati, A. N., Davis, K. A., Lucas, T. H., Litt, B. & Bassett, D. S. Virtual Cortical Resection Reveals Push-Pull Network Control Preceding Seizure Evolution. Neuron 91, 1170–1182 (2016).

16. Khambhati, A. N. et al. Dynamic Network Drivers of Seizure Generation, Propagation and Termination in Human Neocortical Epilepsy. PLOS Comput. Biol. 11, e1004608 (2015).

17. Goodfellow, M. et al. Estimation of brain network ictogenicity predicts outcome from epilepsy surgery. Sci. Rep. 6, 29215 (2016).

18. Panzica, F., Varotto, G., Rotondi, F., Spreafico, R. & Franceschetti, S. Identification of the Epileptogenic Zone from Stereo-EEG Signals: A Connectivity-Graph Theory Approach. Front. Neurol. 4, 175 (2013).

19. Pizzo, F. et al. Epileptogenic networks in nodular heterotopia: A stereoelectroencephalography study. Epilepsia 58, 2112–2123 (2017).

20. Wilke, C., Worrell, G. & He, B. Graph analysis of epileptogenic networks in human partial epilepsy. Epilepsia 52, 84–93 (2011).

21. Cruzat, J. et al. The dynamics of human cognition: Increasing global integration coupled with decreasing segregation found using iEEG. Neuroimage 172, 492–505 (2018).

22. Buch, V. P. et al. Network Brain-Computer Interface (nBCI): An Alternative Approach for Cognitive Prosthetics. Front. Neurosci. 12, (2018).

23. Conrad, E. C. et al. The sensitivity of network statistics to incomplete electrode sampling on intracranial EEG. Netw. Neurosci. 4, 484–506 (2020).

24. Katz, J. S. & Abel, T. J. Stereoelectroencephalography Versus Subdural Electrodes for Localization of the Epileptogenic Zone: What Is the Evidence? Neurotherapeutics 16, 59– 66 (2019).

25. Wagenaar, J. B., Brinkmann, B. H., Ives, Z., Worrell, G. A. & Litt, B. A multimodal platform for cloud-based collaborative research. in 2013 6th International IEEE/EMBS Conference on Neural Engineering (NER) 1386–1389 (IEEE, 2013). doi:10.1109/NER.2013.6696201

26. Kini, L. G., Davis, K. A. & Wagenaar, J. B. Data integration: Combined Imaging and Electrophysiology data in the cloud. Neuroimage 124, 1175–1181 (2016).

27. Azarion, A. A. et al. An open-source automated platform for three-dimensional visualization of subdural electrodes using CT-MRI coregistration. Epilepsia 55, 2028– 2037 (2014).

28. Avants, B., Tustison, N. & Song, G. Advanced Normalization Tools (ANTS). Insight J. 1– 35 (2009).

29. Yushkevich, P. A. et al. User-guided 3D active contour segmentation of anatomical structures: Significantly improved efficiency and reliability. Neuroimage 31, 1116–1128 (2006).

30. Tzourio-Mazoyer, N. et al. Automated Anatomical Labeling of Activations in SPM Using a Macroscopic Anatomical Parcellation of the MNI MRI Single-Subject Brain. Neuroimage 15, 273–289 (2002).

31. Newman, M. E. J. Modularity and community structure in networks. Proc. Natl. Acad. Sci. U. S. A. 103, 8577–8582 (2006).

32. De Meo, P., Ferrara, E., Fiumara, G. & Provetti, A. Generalized Louvain method for community detection in large networks. arXiv (2012).

33. Guimerà, R. & Nunes Amaral, L. A. Cartography of complex networks: modules and universal roles. J. Stat. Mech. Theory Exp. 2005, P02001–1 (2005).

34. Ramaraju, S. et al. Removal of Interictal MEG-Derived Network Hubs Is Associated With Postoperative Seizure Freedom. Front. Neurol. 11, (2020).

35. Wang, Y. et al. Interictal intracranial electroencephalography for predicting surgical success: The importance of space and time. Epilepsia epi.16580 (2020). doi:10.1111/epi.16580

36. Bartolomei, F. et al. Defining epileptogenic networks: Contribution of SEEG and signal analysis. Epilepsia 58, 1131–1147 (2017).

37. Betzel, R. F. et al. Structural, geometric and genetic factors predict interregional brain connectivity patterns probed by electrocorticography. Nat. Biomed. Eng. 1 (2019). doi:10.1038/s41551-019-0404-5

38. Frauscher, B. et al. Atlas of the normal intracranial electroencephalogram: neurophysiological awake activity in different cortical areas. Brain 141, 1130–1144 (2018).

39. Sanz-Garcia, A., Rings, T. & Lehnertz, K. Impact of type of intracranial EEG sensors on link strengths of evolving functional brain networks. Physiol. Meas. 39, (2018).

40. Dam, A. M. Epilepsy and Neuron Loss in the Hippocampus. Epilepsia 21, 617–629 (1980).

41. Lopim, G. M. et al. Relationship between seizure frequency and number of neuronal and non-neuronal cells in the hippocampus throughout the life of rats with epilepsy. Brain Res. 1634, 179–186 (2016).

42. Oldham, S. et al. Consistency and differences between centrality measures across distinct classes of networks. PLoS One 14, e0220061 (2019).

43. Baud, M.O., Kleen, J.K., Mirro, E. A. Multi-day rhythms modulate seizure risk in epilepsy. Nat. Commun. 9, (2018).

