## Supplemental Materials for "Electrocorticography and stereo EEG provide distinct measures of brain connectivity: Implications for network models"

Contents:

Table S1: Extended patient information
Figure S1: Anatomic distribution of electrodes is similar in ECoG and SEEG
Figure S2: Network connectivity in alternate frequency bands
Figure S3: Distinguishability of resected versus spared tissue in alternate frequency bands

| Portal ID | Surg. | Type. | Lat. | Les. | Age | Sex | Total Node | Depth Node | Total Res. | Depth Res. |
|---|---|---|---|---|---|---|---|---|---|---|
| HUP65_phaseII | RES | ECoG | R | LES | 36 | M | 64 | 0 | 15 | 0 |
| HUP68_phaseII | RES | ECoG | R | NL | 28 | F | 83 | 0 | 24 | 0 |
| HUP74_phaseII | RES | ECoG | L | LES | 25 | F | 108 | 17 | 53 | 17 |
| HUP78_phaseII | RES | ECoG | L | LES | 54 | M | 97 | 0 | 20 | 0 |
| HUP82_phaseII | RES | ECoG | R | LES | 56 | F | 83 | 5 | 40 | 3 |
| HUP88_phaseII | RES | ECoG | L | LES | 35 | F | 53 | 7 | 7 | 0 |
| HUP89_phaseII | RES | ECoG | R | LES | 29 | M | 95 | 7 | 9 | 5 |
| HUP094_phaseII | RES | ECoG | R | NL | 48 | F | 81 | 14 | 2 | 0 |
| HUP097_phaseII | RES | ECoG | L | NL | 39 | F | 91 | 7 | 15 | 1 |
| HUP099_phaseII_D01 | RES | ECoG | R | LES | 20 | F | 105 | 12 | 28 | 6 |
| HUP105_phaseII | RES | ECoG | R | LES | 39 | M | 54 | 7 | 4 | 0 |
| HUP106_phaseII | RES | ECoG | L | NL | 45 | F | 114 | 14 | 9 | 6 |
| HUP107_phaseII | RES | ECoG | R | NL | 36 | M | 114 | 17 | 22 | 8 |
| HUP111_phaseII_D02 | RES | ECoG | R | NL | 40 | F | 100 | 11 | 6 | 3 |
| HUP116_phaseII | ABL | SEEG | R | LES | 59 | F | 34 | 34 | 5 | 5 |
| HUP117_phaseII | RES | SEEG | L | LES | 39 | M | 29 | 29 | 3 | 3 |
| HUP125_phaseII_D04 | ABL | ECoG | L | NL | 57 | M | 108 | 20 | 8 | 8 |
| HUP126_phaseII_D01 | ABL | ECoG | L | NL | 26 | F | 123 | 37 | 9 | 9 |
| HUP138_phaseII | ABL | SEEG | L | LES | 38 | M | 71 | 71 | 3 | 3 |
| HUP140_phaseII_D02 | ABL | SEEG | L | NL | 47 | F | 53 | 53 | 5 | 5 |
| HUP144_phaseII | RES | SEEG | R | LES | 31 | M | 86 | 86 | 15 | 15 |
| HUP146_phaseII | RES | SEEG | R | NL | 16 | M | 86 | 86 | 7 | 7 |
| HUP148_phaseII_D02 | ABL | SEEG | L | LES | 23 | M | 69 | 69 | 7 | 7 |
| HUP157_phaseII | ABL | SEEG | L | NL | 25 | M | 116 | 116 | 6 | 6 |
| HUP160_phaseII | RES | SEEG | R | NL | 45 | F | 65 | 65 | 11 | 11 |
| HUP164_phaseII | ABL | SEEG | L | LES | 34 | F | 131 | 131 | 3 | 3 |
| HUP165_phaseII | ABL | SEEG | R | NL | 21 | F | 151 | 151 | 10 | 10 |
| HUP173_phaseII | RES | SEEG | R | LES | 24 | F | 87 | 87 | 17 | 17 |
| HUP177_phaseII | RES | SEEG | R | NL | 42 | F | 139 | 139 | 16 | 16 |
| HUP181_phaseII_D02 | ABL | SEEG | L | LES | 31 | F | 103 | 103 | 6 | 6 |
| HUP185_phaseII | ABL | SEEG | L | LES | 38 | M | 93 | 93 | 9 | 9 |
| HUP187_phaseII | ABL | SEEG | R | NL | 25 | M | 77 | 77 | 7 | 7 |
| HUP190_phaseII | RES | SEEG | L | NL | 25 | M | 117 | 117 | 25 | 25 |

**Table S1: Extended patient information.** Column 1) ieeg.org portal ID where full-length records are freely available & searchable. Column 2) Surgery type – RES: resection, ABL: laser ablation. Column 3) Laterality (Right/Left). Column 4) Pre-op lesion status – LES: lesional, NL: non—lesional. Column 5) Sex (Male/Female). Column 6) Total number of nodes in grey matter (GM). Column 7) Total number of GM nodes sampled by depth electrodes. Column 8) Total number of resected GM nodes. Column 9) Total number of resected GM nodes sampled by depth electrodes.

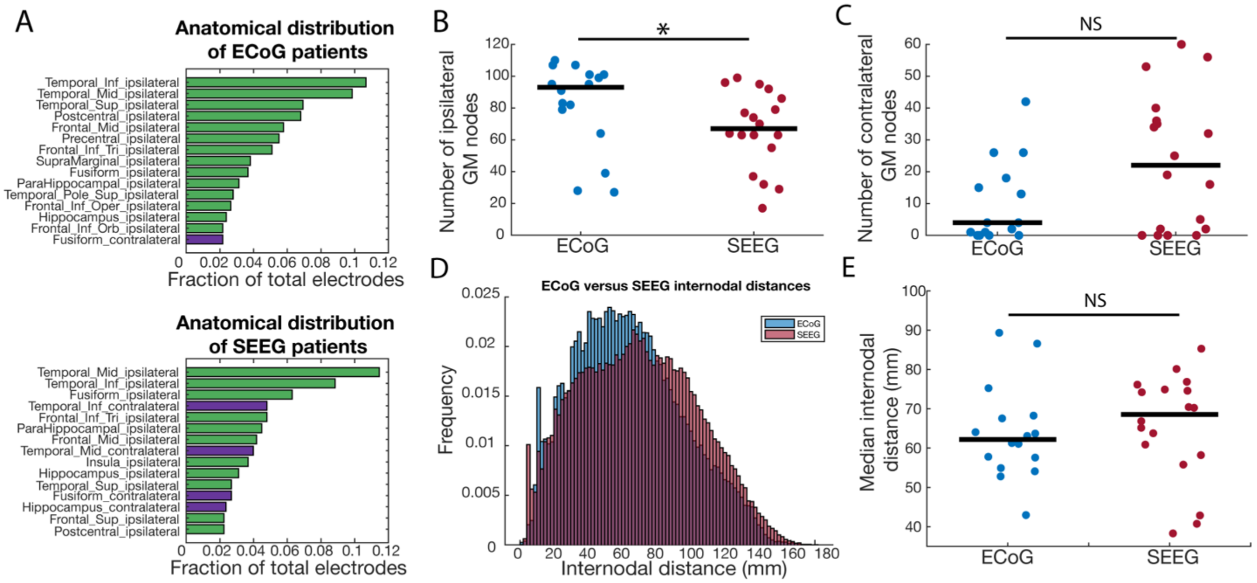


**Figure S1. Anatomic distribution of electrodes is similar in ECoG and SEEG:** (A) The top 15 ranked AAL regions in terms of total number of electrodes across patients. Green: ipsilateral, Purple: contralateral. Abbreviations: Mid: middle, Inf: inferior, Sup: superior, Tri: pars triangularis, Oper: pars opercularis, Orb: pars orbitalis. (B) The number of grey matter nodes ipsilateral to the resection zone was higher in ECoG (median: 93) versus SEEG (median: 64), (rank-sum test, $p = 0.022$). (C) Comparing the number of grey matter nodes contralateral to the resection zone between ECoG (median: 4), and SEEG (median: 25) did not reach statistical significance (rank-sum test, $p = 0.057$). (D) Distribution histogram of internodal distances in ECoG (blue) and SEEG (red). (E) The median internodal distance in each patient was not significantly different between ECoG (median: 62.2) versus SEEG (median: 70.2), (rank-sum test, $p = 0.24$).

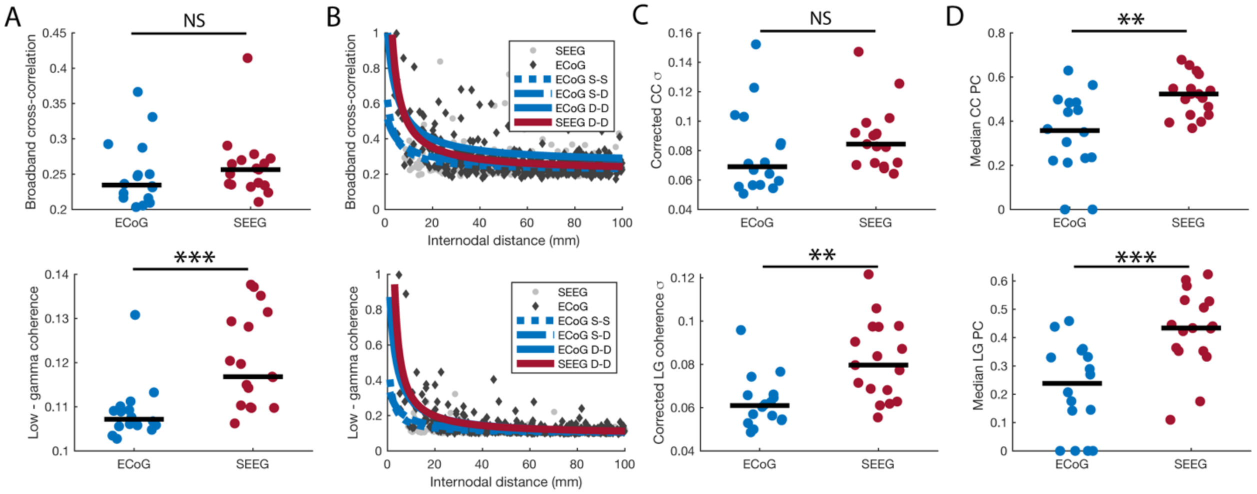


**Figure S2. Network connectivity in alternate frequency bands:** Data for broadband cross-correlation (CC) are on the top row, and low-gamma (LG) coherence is on the bottom row. (A) Median connectivity values were not significantly different between ECoG and SEEG for broadband cross-correlation (rank-sum test p = 0.17), but SEEG had higher low-gamma coherence (rank-sum test p = 1.9e-4). (B) We fit a nonlinear regression model to ECoG surface – surface (dotted blue line), surface – depth (dashed blue line), and depth – depth connections (solid blue line), as well as SEEG depth – depth connections (solid red line). (C) After correcting for internodal distance, the standard deviation of edge weights remained higher in SEEG versus ECoG for low-gamma coherence (LG rank-sum test p = 0.0013) but not broadband cross-correlation (CC rank-sum test p = 0.10). (D) After correcting for internodal distance, the median participation coefficient remained higher in SEEG versus ECoG (CC rank-sum test p = 0.0026, LG rank-sum test p = 8.5e-4). Abbreviations: NS = not significant, S-S: surface – surface, S-D: surface – depth, D-D: depth – depth, SD: standard deviation, ** = p < 0.01, *** = p < 0.001.

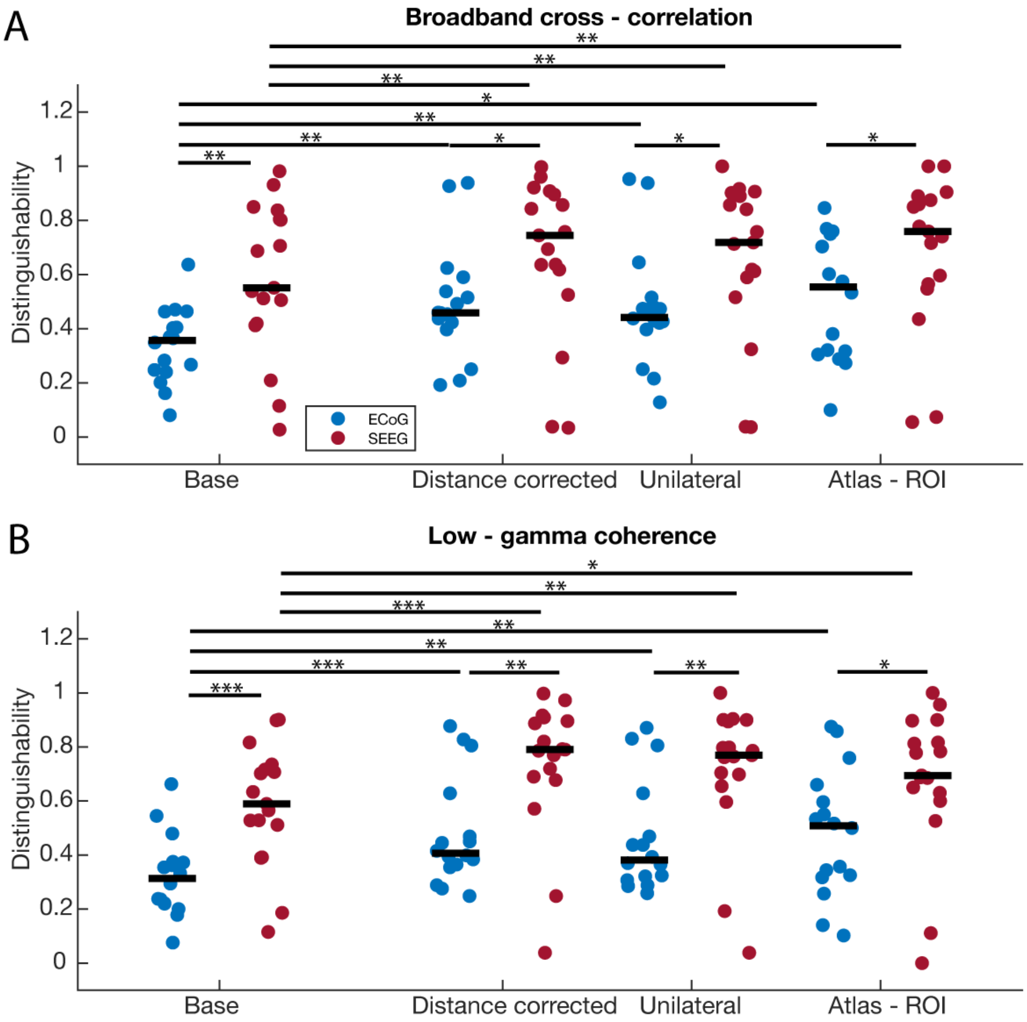


**Figure S3. Distinguishability of resected versus spared tissue in alternate frequency bands:** We computed distinguishability for the different networks derived from ECoG and SEEG (Figure 4A), including only grey matter (GM), distance corrected (DC), networks with contralateral nodes eliminated (UL), and networks with atlas-level ROI (AR). (A) For broadband cross-correlation in each condition, SEEG networks had a higher $D_{rs}$ value than ECoG. Each condition in ECoG and SEEG also had a higher $D_{rs}$ value than not accounting for internodal distance. GM ECoG vs GM SEEG: (rank-sum test $p = 0.0042$), DC ECoG vs DC SEEG: (rank-sum test $p = 0.024$), UL ECoG vs UL SEEG: (rank-sum test $p = 0.029$), AR ECoG vs AR SEEG: (rank-sum test $p = 0.029$), DR/UL/MR ECoG vs GM ECoG: (sign-rank test p = 0.0023/0.0072/0.0131), DC/UL/AR SEEG vs GM SEEG: (sign-rank test p = 0.0012/0.0019/0.0075). (B) For low – gamma coherence: SEEG networks had a higher $D_{rs}$ value than ECoG. Each condition in ECoG and SEEG also had a higher $D_{rs}$ value than not accounting for internodal distance. GM ECoG vs GM SEEG: (rank-sum test $p = 9.8e-4$), DC ECoG vs DC SEEG: (rank-sum test $p = 0.0059$), UL ECoG vs UL SEEG: (rank-sum test $p = 0.0090$), AR ECoG vs AR SEEG: (rank-sum test $p = 0.014$). DC/UL/AR ECoG vs GM ECoG: (sign-rank test p = 0.0009/0.0027/0.0052), DC/UL/AR SEEG vs GM SEEG: (sign-rank test p = 0.0007/0.0010/0.0245).
